# CLINICAL EFFICACY OF PLASMID ENCODING P62/SQSTM1 (ELENAGEN) IN COMBINATION WITH GEMCITABINE IN PATIENTS WITH PLATINUM-RESISTANT OVARIAN CANCER

**DOI:** 10.1101/2023.08.08.23292616

**Authors:** Sergei Krasny, Yauheni Baranau, Sergey Polyakov, Ekaterina Zharkova, Olga Streltsova, Aliona Filimonava, Volha Siarheyeva, Sviatlana Kazlouskaya, Anton Khorau, Vladimir Gabai, Alexander Shneider

## Abstract

**Purpose:** The purpose of this clinical study is to evaluate safety and efficacy of ELENAGEN, a novel anticancer therapeutics (plasmid DNA encoding p62/SQSTM1) protein, as an adjuvant to chemotherapy with Gemcitabin (GEM) in patients with advanced platinum-resistant ovarian cancer.

**Patients and Methods:** This was a prospective randomized multi-center study with two arms. Gemcitabine 1000 mg/m^2^ days 1,8 every 3 weeks) was administered in both arms: In the Chemo arm (n = 20) GEM was the only treatment, and in the ELENAGEN arm (n = 20) GEM was supplemented with ELENAGEN (2.5 mg i.m. weekly). The primary endpoint was progression-free survival (PFS), and the secondary endpoint was safety. Antitumor activity was assessed by RECIST 1.1 criteria. Safety was assessed on the basis of adverse events (AEs) and serious AEs (SAEs) according to NCI CTCAE version 5.0.

**Results:** To data cut-off, the median follow-up was 13.8 months. There were no SAE -related to ELENAGEN treatment. The median progression-free survival (PFS) was 2.8 and 7.2 mo in Chemo and ELENAGEN arms respectively (p Log-Rank = 0.03). Noteworthy, at the time of cut-off, 9 patients (45%) in Elenagen arm did not progress with the longest PFS recorded so far is 24 months. Subgroup analysis of patients in both arms demonstrated high efficacy of Elenagen in the patients with worse prognosis: high pretreatment levels of CA125, progression after only one line of chemotherapy, and peritoneal effusion.

**Conclusions:** Addition of ELENAGEN to Gemcitabine is effective in patients with ovarian cancer, including those with a worse prognosis.

## Introduction

About 20 000 new cases of ovarian cancer (OC) is diagnosed in the US every year; and it is the most lethal gynecologic malignancy with an overall 5-year survival of only 50% (https://cancerstatisticscenter.cancer.org/#!/cancer-site/Ovary). This high lethality occurs due to the fact that the majority of patients are diagnosed at an advanced stage, and, following front-line therapy, eventually tumors become chemoresistant. While 80% of patients can achieve remission after initial treatment, a majority will relapse within 1.5 year (1). Platinum-based chemotherapy, usually in combination with taxanes, remains a cornerstone of systemic therapy for advanced and recurrent ovarian cancer. However, recurrent OC remains difficult to treat due to chemotherapy resistance. In recent years, the development of anti-angiogenic and poly ADP-ribose polymerase I (PARP) inhibitors have modestly improved patient progression-free survival (PFS) (2) (3,4). Anti-angiogenic agents (e.g. bevacizumab) have demonstrated clear antitumor activity by delaying recurrence and progression in patients with ovarian cancer, but translating that benefit to prolonged overall survival remains challenging (2). Thus, a novel OC therapeutics to improve long-term outcomes are urgently needed.

Recently, immunotherapy of cancer, especially with immune-checkpoint inhibitors (ICI), emerged as a novel treatment option for a number of solid tumors, and it was also tested in several clinical trials with OC (5). However, unlike other tumor types, the results of the trials were not encouraging. For instance, in patients with platinum-resistant OC, comparing with standard chemotherapy with gemcitabine (GEM) or pegylated lyposomal doxorubicin (PLD), PFS with one of ICI nivolumab (anti-PDL1 Antibody) was only 2.0 mo vs 3.8 mo with GEM or PLD, and OS was 10.1 vs 12.1 mo (6). Also, a grade 3 related adverse events (AE) occurred in 33% of patients in the nivolumab group (6). In JAVELIN Ovarian 200 phase III trial of 566 patients with platinum-resistant OC, addition of another anti-PD-L1 Antibody, Avelumab, to standard PLD treatment did not statistically increase PFS (3.7 vs 3.5) or OS (15.7 vs 13.1) (7). Furthermore, serious treatment-related adverse events occurred in 18% patients in the combination group, comparing with 11% in the PLD only group (7). Thus, at present, application of ICI in the treatment of platinum-resistant OC does not look encouraging.

We have recently developed a novel anticancer therapeutics ELENAGEN based on plasmid DNA encoding p62 (SQSTM1) protein (8). p62 is multifunctional protein which participates in selective autophagy, signal transduction, inflammatory response and other processes (9). p62 can be a good target for anticancer vaccine since its levels are elevated in almost all human tumors tested so far and it increases when tumors progress (see ref ((10,11) for review). While p62 is dispensable for normal cells, tumors require p62 for growth and metastases (10).

Importantly, p62 levels are also increased in OC and it associates with poor prognosis and platinum resistance making p62 a good target for in immune response (12,13).

Besides evoking an immune response, ELENAGEN can also alleviate a chronic inflammation by suppressing generation of proinflammatory cytokines such as TNF, IL-1, IL-6 in different rodent disease models (14) (15). In contrast to acute inflammation that is beneficial for immune response to microbes and cancer cells, intratumoral chronic inflammation is detrimental since it disables immune cells thus suppressing anti-tumor immunity (see ref (16) for review). Since most chemo-therapeutics (at least partially) engage the immune system as part of their anti-tumoral mechanism of action, chronic inflammation decreases sensitivity to chemotherapy and prevents drugs delivery to tumors (17).

The tumor milieu of OC is also enriched with a broad spectrum of pro-inflammatory cytokines and chemokines. In particular, several of these cytokines such as tumor necrosis factor (TNF)-a, interleukin (IL)-1β and IL-6 produced by tumor itself or/and activated immune cells, besides stimulating cancer cell growth, also influenced disease progression and prognosis by reducing responsiveness to chemotherapy (18). For instance, elevated levels of proinflammartory cytokine IL-6 in serum or ascites of OC patients correlated with chemoresistance, in particular, platinum resistance (18). Since ELENAGEN can alleviate chronic inflammation, it may render tumor cells more susceptible to chemotherapy and the immune attack. Therefore, two mechanisms of ELENAGEN action, as anticancer vaccine and anti-inflammatory drug, are complimentary and can make it in combination with chemotherapeutic agents a unique anti-cancer therapeutics for treatment of OC.

We have previously conducted a phase I/IIa clinical trial of ELENAGEN used as a mono-therapy (19). In that study, ELENAGEN showed promise in treating patients with advanced disease for which all standard methods of treatment were exhausted. For example, progression of OC was stopped for three or more months in 4 out of 6 patients. Importantly, in contrast to ICI (see above), AE during ELENAGEN treatment were only of Grade 1 and no severe AE were observed (19). These data encouraged us to conduct a current clinical study of ELENAGEN with platinum-resistant OC.

## PATIENTS AND METHODS

### Study design and patients

Eligible patients were ≥18 years old; had measurable ovarian cancer per RECIST 1.1 criterion that had progressed <6 months after completion of platinum-based therapy; an Eastern Cooperative Oncology Group performance status (ECOG PS) of 0 or 1; adequate hematologic and organ functions (see Suppl 1 for inclusion/exclusion criteria).

The study was conducted in accordance with the Declaration of Helsinki and International Conference on Harmonization E6 guidelines, and was approved by the relevant institutional review boards of participating research centers.

All patients provided written informed consent and agreed to provide archival and/or freshly collected tumor tissue and blood samples

## TREATMENT

This was a prospective randomized study with two arms. Chemotherapy (GEM) 1000 mg/m^2^ days 1,8 every 3 weeks) was administered in both arms. In the Chemo arm (n = 20) it was the only treatment, and in the ELENAGEN arm (n = 20) the same chemotherapy was supplemented with ELENAGEN (2.5 mg i.m. weekly).

## ASSESSMENT AND ENDPOINTS

In Safety Analysis Set and in Efficacy-Evaluable Set all patients who received ≥ 1 dose (n = 20 patients in each arm Chemo and ELENAGEN arms) were included. Safety was assessed on the basis of adverse events (AEs) and serious AEs (SAEs) according to NCI Common Terminology Criteria for Adverse Events version 4.0.

Antitumor activity was assessed by investigator according to RECIST 1.1 criteria. Evaluation of the therapeutic effect was carried out by computer tomography (CT) 19-20 days after the completion of each 2nd course of chemotherapy (before 3-,5-, 7-, 9, 11-th courses, on a visit of completion of treatment, and, if necessary, on follow-up visits). In the absence of tumor progression during the course of treatments, the effectiveness of treatment by CT was evaluated every 6 weeks. PFS was defined as the time from study treatment initiation to the first occurrence of documented disease progression or death from any cause during the study, whichever occurred first.

## STATISTICAL ANALYSES

Tumor response was evaluated according to the RECIST ver. 1.1. Response assessment were performed every 3 courses of chemotherapy. PFS was defined as the time from date of randomization to the first event of disease progression and assessed using Kaplan-Mayer method. PFS in the two treatment arms was compared using an unstratified two-sided log-rank test. A P _D_<_D_0.05 was considered as statistical significance. For the subgroup analyses proportional COX regression model were used.

## RESULTS

Forty patients with platinum-resistant ovarian cancer were enrolled in the study with 20 patients in each arm (GEM or GEM+ELENAGEN). To the date of cutoff, In GEM (Chemo) arm all patients have been progressed, and in ELENAGEN arm 9 patients (45%) continued follow up.

Patients characteristics are summarized in Table 1. At diagnosis, most patients in both groups (75-85%) had histologically high grade serous adenocarcinoma. More than half of patients in both groups (55-65%) had one line of platinum chemotherapy before progression, and platinum-free interval 3-6 months (60-65%). Also majority of patients in both groups had high levels of CA125 oncomarker (75-80%), as well as metastases in peritoneum (75-85%) and elsewhere (Table 1).

**Table 1.**
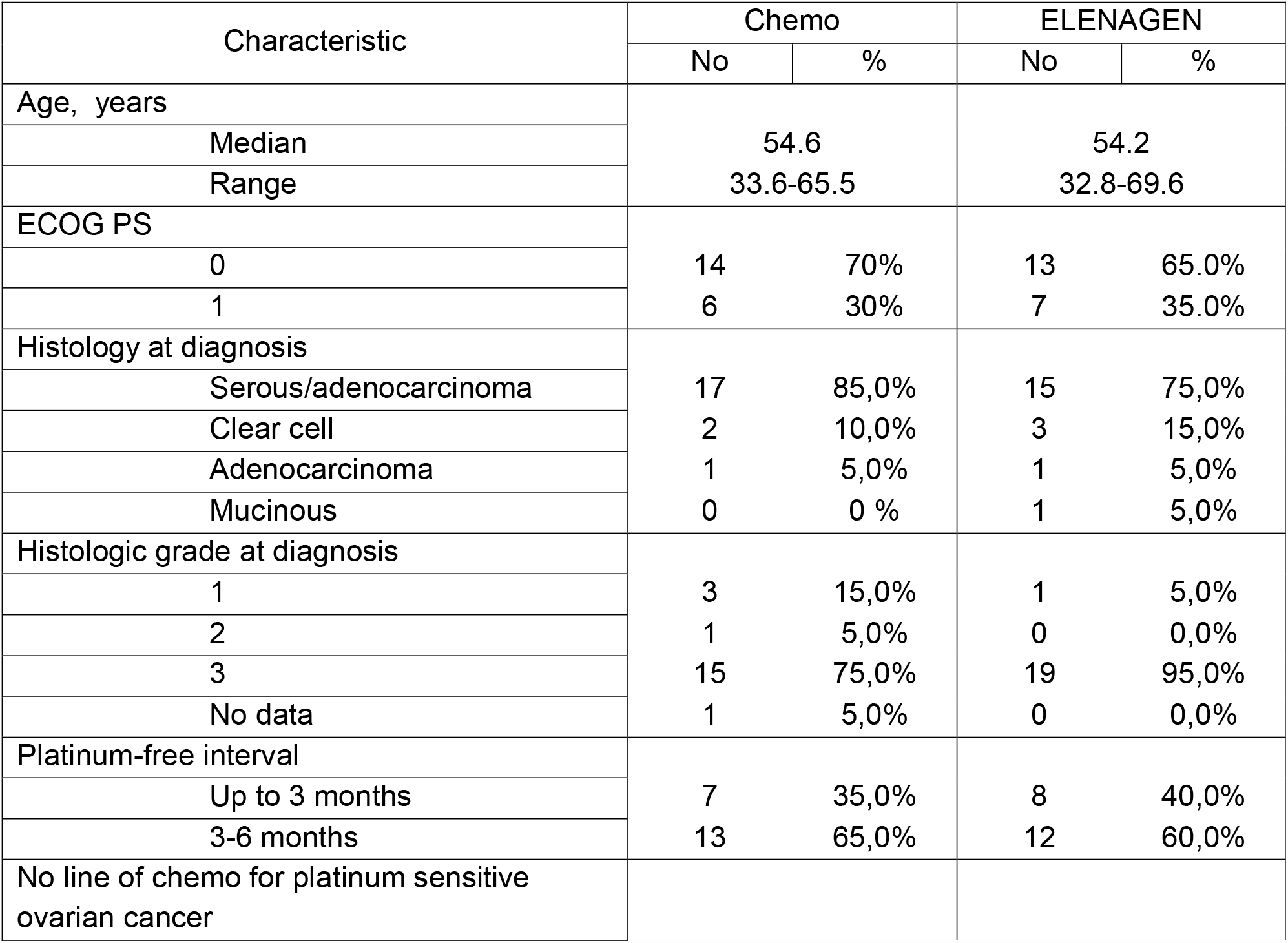
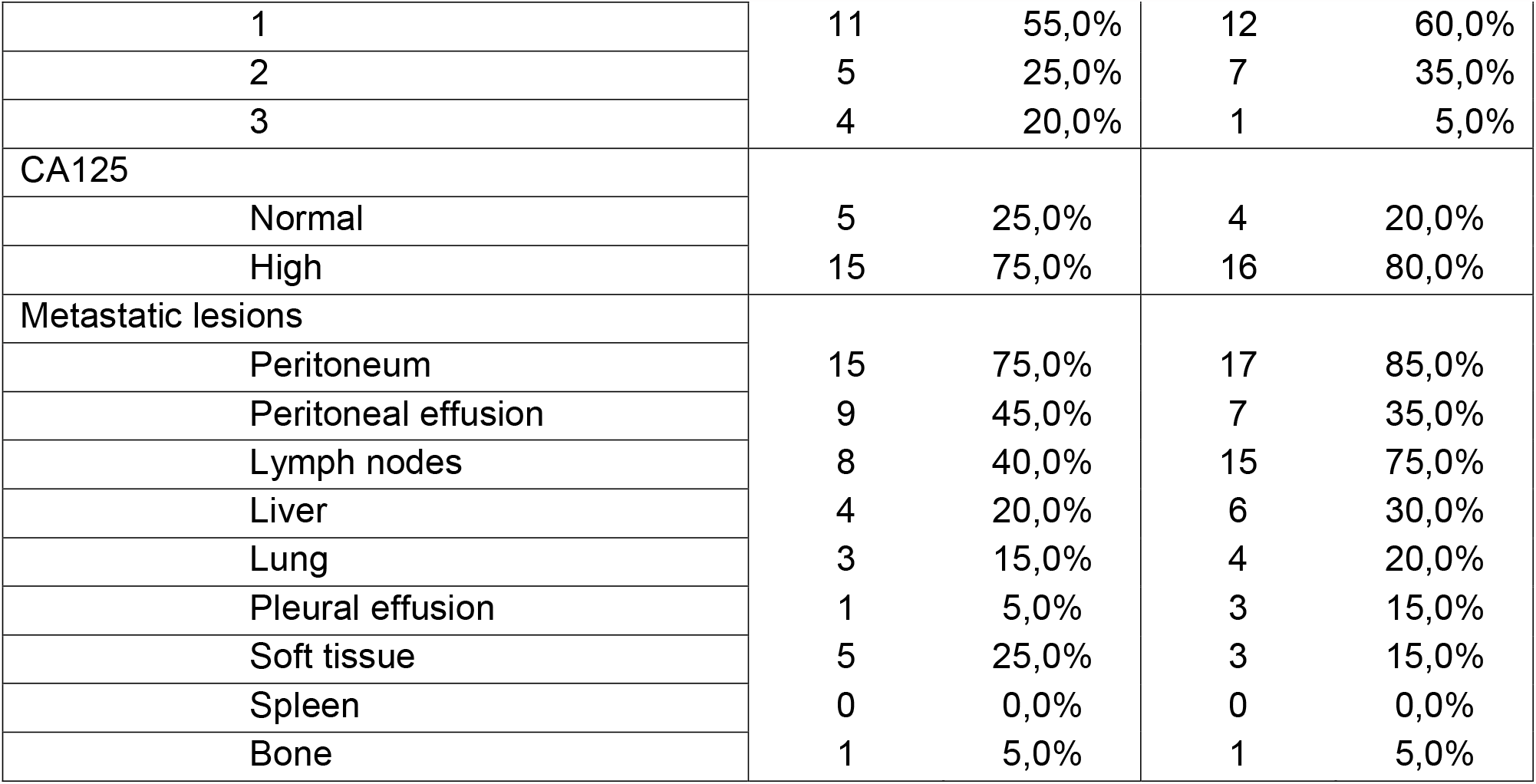
Baseline Patient Characteristics.

### Safety

The safety was assessed in all 40 patients. During the study period, one death was registered in Plasmid arm without any evidence of disease progression within 2 months after randomization, and its possible cause is venous embolism. Although autopsy was not performed and the final diagnosis has not determined, this adverse event was counted as thrombosis and unrelated to the disease. One patient in Plasmid arm underwent surgery due to intestinal obstruction within one month after randomization, and the subsequent cycle of the treatment was delayed for three weeks. After recovery from the surgery the patient continued treatment without evidence of progression.

The majority of adverse events in Chemo and Plasmid arms were related to GEM side effects and manifested, as expected, by hematological toxicity: neutropenia, thrombocytopenia, and anemia. No cases of febrile neutropenia or other life-threatening complications which required hospitalization occurred. A fraction of adverse events was related to the disease process: intestinal obstruction, and increase in creatinine/urea levels due to retroperitoneal lymph nodes compression. Only skin rash, itching and redness at the injection site were considered as related to Plasmid administration. At the same time the number of Adverse events with Grade <= 3 and AE of special interest (potentially related to plasmid administration) did not significantly differ between the groups (Suppl Table1).

A slight increase in the number of hematological adverse events in the plasmid arm was apparently related to the longer GEM exposure due to higher efficacy and increasing in PFS of combination GEM and Plasmid over Gemcitabine alone (see Fig. 1 below).

**Fig. 1.**
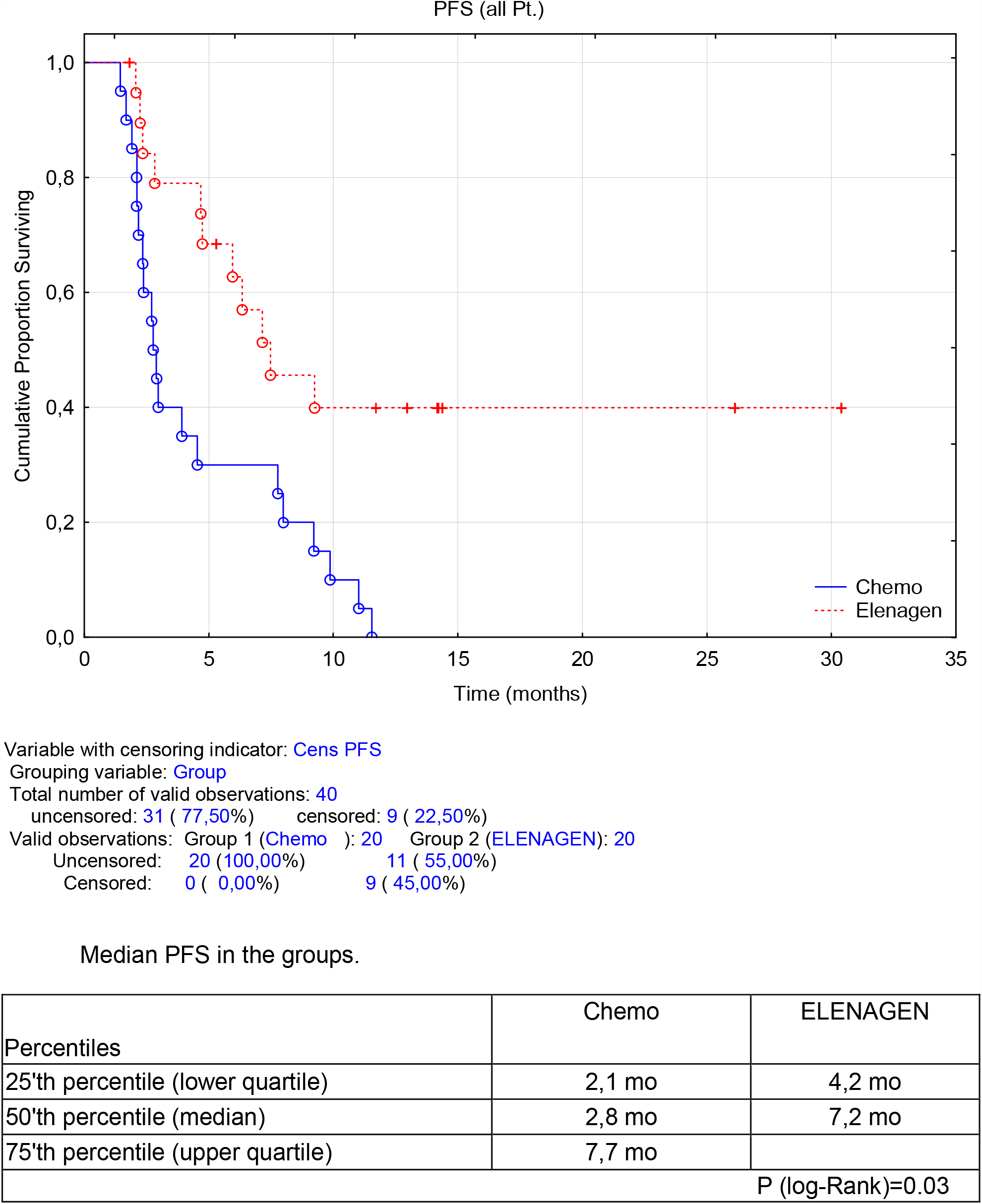
Progression-free survival of patients treated with Chemo+ELENAGEN, or Chemo only

### Efficacy

To data cut-off, the median follow-up was 13.8 months in Efficacy-Evaluable Set. The tumor response was assessed according to the RECIST 1.1 criteria. No complete responses were observed in either group. The objective response rate was higher in the ELENAGEN arm: partial response (PR) 5.9% and 26.7%, stable disease (SD) 35.3% and 53.3%, disease progression 58.8% and 20.0% in Chemo and ELENAGEN arm respectively. Totally, disease control rate (PR and SD) was significantly higher in ELENAGEN arm (80.0% vs 41.2% in Chemo and ELENAGEN arm, respectively, p = 0,001)). One patient in the ELENAGEN arm was able to undergo a complete cytoreduction with no evidence of disease progression.

The median progression-free survival (PFS) was 2.8 and 7.2 mo in Chemo and ELENAGEN arms respectively (p Log-Rank = 0.03) (Fig. 1) For the lower 25^th^ percentile (lower quartile) these numbers were 2.1 vs. 4.2 months respectively, while for the upper quartile (75^th^ percentile) it was only possible to determine for the chemotherapy group alone, 7.7 months. Noteworthy, at the time of cut-off, 9 patients (45%) in ELENAGEN arm did not progress with the longest PFS recorded so far is 24 months.

### Subgroup Analysis

We assessed efficacy of ELENAGEN in subgroups with different basic characteristics Due to randomization, the number of the patients in each of these subgroup of Chemo and ELENAGEN arms were almost equal. When we analyzed response to ELENAGEN in the subroups with higher than normal levels of CA125 oncomarker (>35 U/ml), the difference between Chemo and ELENAGEN arms (15 and 16 patients in each arm with high levels of CA125) was statistically more pronounced indicating that the more severe patients with high levels of CA125 before treatment were very responsive to ELENAGEN supplementing Chemo (p=0.01) (Fig.2).

**Fig. 2.**
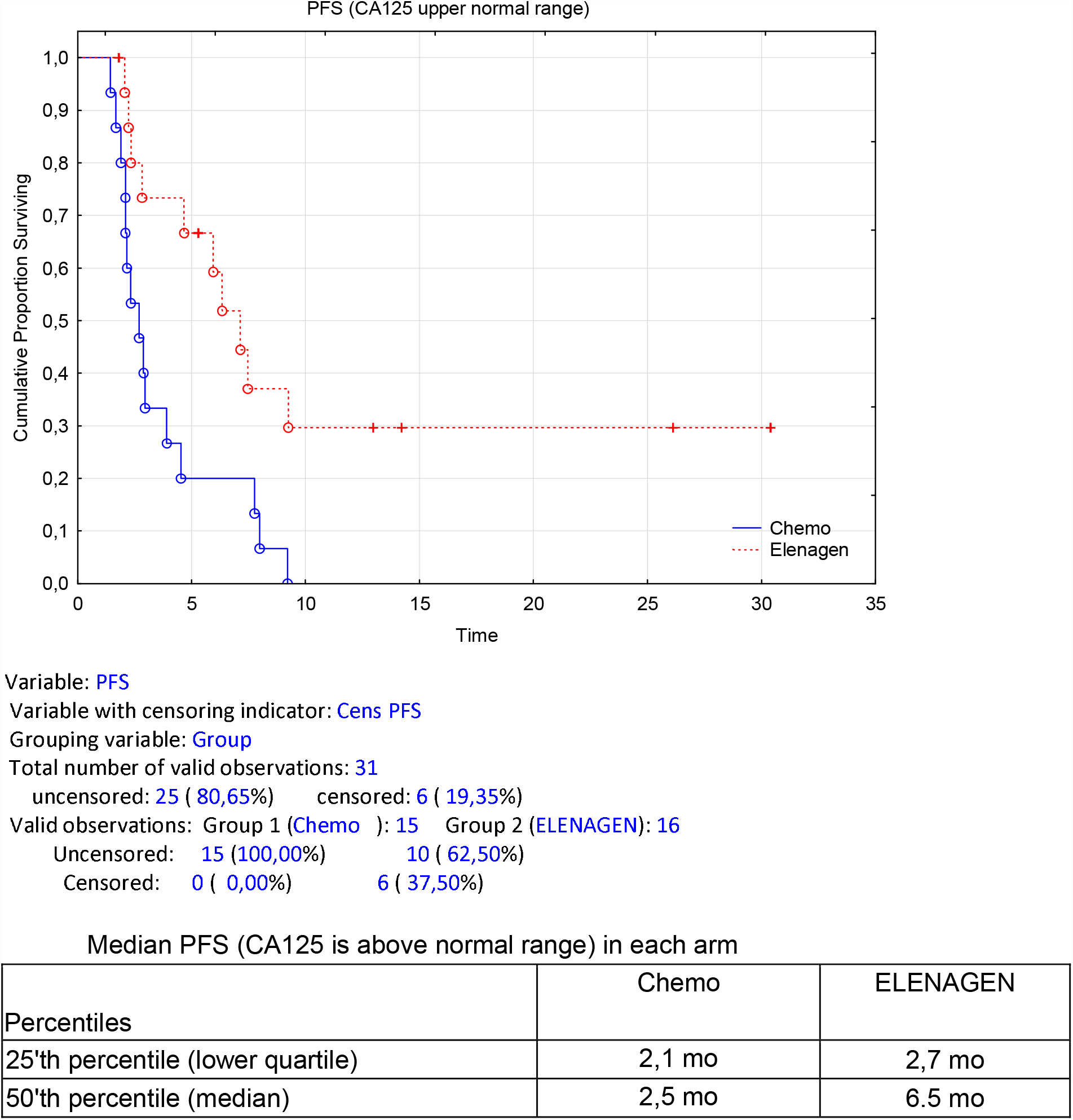
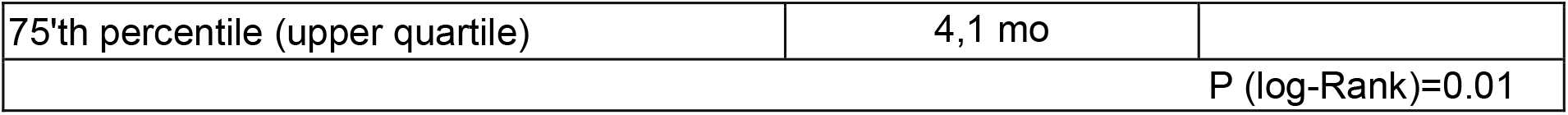
Progression-free survival of patients treated with Chemo+ELENAGEN, or Chemo only in subgroups of patients with higher than normal levels of CA125 (>35 U/ml)

Next we analyzed the responses to the treatments in the subgroup of patients who progressed after only one line of platinum therapy. Comparison of such patients (11 in Chemo group and 12 in ELENAGEN group) demonstrated highly significant response in ELENAGEN arm comparing to Chemo group (median PFS= 2.3 mo vs 7.1 mo in Chemo and ELENAGEN arms, respectively, p<0.01). These results demonstrate ELENAGEN enhances Chemo response even that even in more severe patients who progressed after only one line of chemotherapy, it is still sensitive to ELENAGEN treatment (Fig.3).

**Fig. 3.**
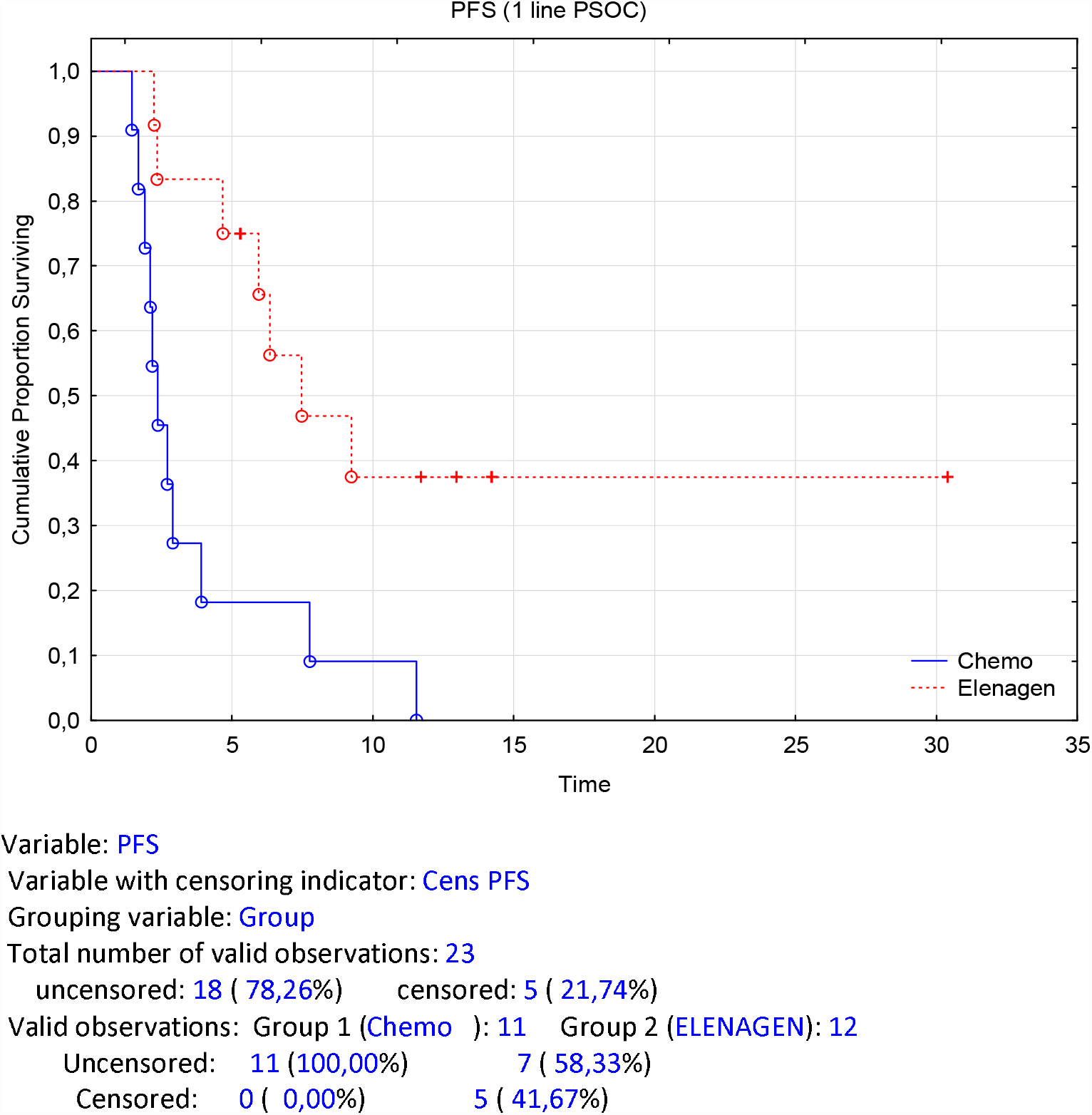
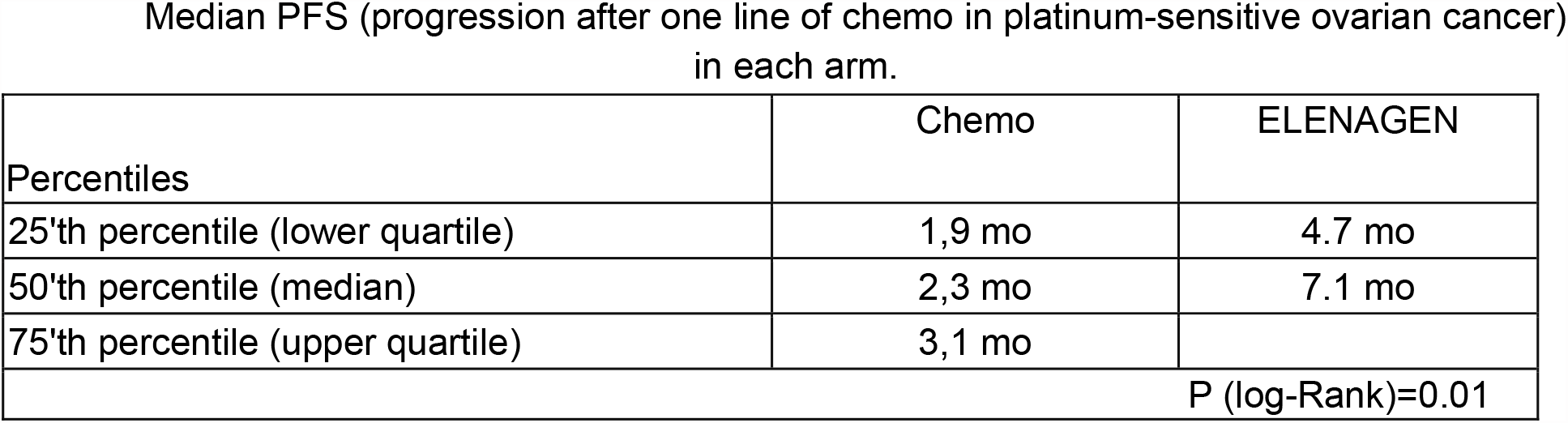
Progression-free survival of patients treated with Chemo+ELENAGEN, or Chemo only in subgroups of patients who progressed after one line of chemotherapy

Finally, we assessed treatment responses in the subgroups of the patients who had peritoneal effusion (ascites) before treatment. Whereas in Chemo group median PFS was 2.4 mo, in ELENAGEN group it increased more than 3-times, to 7.6 mo demonstrating that these patients are also more sensitive to ELENAGEN plus Chemo treatment with p=0.008 (Suppl Fig.1).

## DISCUSSION

Platinum-resistant OC, even if treated with a standard therapy such a gemcitabine, PLD, paclitaxel, and topotecan have a dismal prognosis: medium PFS of 3-4 mo and OS is 12 months (20,21). Therefore, a more effective therapy for this form of OC is urgently needed. Despite success of immunotherapy of immune checkpoint inhibitors (ICI) in some tumors (22)), such combination of ICI with chemotherapy in OC so far was not successful, and this treatment was quite toxic (5) (7) (see Introduction). Thus, at present, application of ICI in the treatment of platinum-resistant OC does not look encouraging.

Elevated levels of IL-6 in serum or ascites of OC patients correlated with chemoresistance, (18), and higher ascites levels of IL-6 and TNF predicts worse PFS in patients with OC (23). Elenagen was shown to decrease a chronic inflammation (24), that may promote effect of chemotherapy in OC (17). In the current study we tested if ELENAGEN would enhance standard anti-OC chemotherapy, GEM, known to stimulate anti-cancer immunity. Reciprocally, GEM deplete immunosuppressive MDSC (25,26) which may enhance anti-cancer effect of Elenagen as DNA vaccine.

Here demonstrated that addition of our novel plasmid drug ELENAGEN to a standard chemotherapy regimen with GEM had a profound effect on PFS increasing it from 2.8 mo to 7.2 mo. Importantly, no signs of increased toxicity of this combined treatment comparing to GEM alone was found. Remarkably, ELENAGEN combination with GEM was also effective in patients with dismal prognosis: with high pretreatment levels of CA125, progression after only one line of chemotherapy, and peritoneal effusion. For instance, recent meta-analysis of data from more than 10 000 patients demonstrated that the increased serum level of CA-125 before treatment was correlated with poor progression-free (HR=1.59, 95%CI=1.44∼1.76, p<0.001) and overall survival (HR=1.62, 95% CI=1.270-2.060, p<0.001) (27).

In conclusion, addition of ELENAGEN to Gemcitabine is effective in patients with ovarian cancer, including those with a worse prognosis. Future studies of ELENAGEN with different chemo-or radiotherapy regimens are warranted.

## Data Availability

All data produced in the present study are available upon reasonable request to the authors

