## Supplement for "CLINICAL EFFICACY OF PLASMID ENCODING P62/SQSTM1 (ELENAGEN) IN COMBINATION WITH GEMCITABINE IN PATIENTS WITH PLATINUM-RESISTANT OVARIAN CANCER"

Supplemental data

**Criteria for inclusions/exclusions**

• The patient is 18-65 years old.

• Written informed consent of the patient to participate in clinical trials.

• Presence of histologically confirmed ovarian cancer.

• The return of the disease occurred less than 6 months after the last administration of platinum.

• Presence of measurable tumor lesions according to RECIST 1.1 criteria.

• Functional status according to ECOG scale is 0-2.

• Life expectancy of at least 6 months.

• Adequate function of the organs as determined by the following criteria:

a. Absolute number of neutrophils (ANN) ≥1500/mm3 (≥1.5 × 109/l);

b. Platelet count ≥100,000/mm3 (IU: ≥100 × 109/l).

c. Hemoglobin level ≥ 9.0 g/dL determined by analysis performed at least 2 weeks after the last hemotransfusion;

d. The level of AST and ALT in blood serum not exceeding more than 3 times the upper limit of the norm.

e. The level of serum bilirubin not exceeding more than 1.5 times the upper limit of normal.

f. Serum Creatinine ≤ 1.5 mg/dL.

• The ability of the patient to follow the directions of the research physician and follow the study regimen.

**Criteria by which patients are not included in the study**

• Platinum-sensitive relapse of ovarian cancer (disease recurrence occurred more than 6 months after the last administration of platinum drugs).

• Presence of serious diseases or health conditions:

a. Other malignancies, with the exception of malignancies treated more than 5 years ago without signs of return of the disease.

b. Brain metastases or leptomeningeal metastases.

c. Active infection (e.g. fever ≥38 °C), including active or unresolved pneumonia/pneumonitis.

d. Uncontrolled diabetes mellitus.

e. Myocardial Infarction in the last 12 months, severe/unstable angina, clinically manifested heart failure class III-IV according to the classification of the New York Cardiology Association (NYCA).

f. Gastrointestinal bleeding within the last 2 weeks.

g. Human immunodeficiency virus (HIV), chronic or acute hepatitis B or hepatitis C.

h. Patients with autoimmune disorders or organ transplantation who require immunosuppressive therapy.

I. Mental illness that may increase the risk associated with participation in the study or taking the study drug or that may affect the interpretation of the results of the study.

K. Polyallergy, bronchial asthma (including aspirin) in history.

• Major surgery during the previous 4 weeks (complete wound healing).

• Previous chemotherapeutic treatment of the patient according to the scheme **GEMCITABINE**

• Radiotherapy with extended field radiation within the previous 4 weeks or radiotherapy with a limited field radiation within the previous 2 weeks.

**Criteria for exclusion (exit) from the study**

- Individual intolerance to the drug.

-The patient's desire to stop the study.

- Serious adverse events occurring in the patient during the study.

-Violation of conditions of the trial of the drug by patient (non-compliance).

Suppl Table 1. Adverse events Grade <= 3 and AE of special interest

| Adverse event | Chemo arm | | Plasmid arm | |
| --- | --- | --- | --- | --- |
|  | No | % | No | % |
| Neutropenia | 4 | 20,0% | 7 | 35,0% |
| Thrombocytopenia | 2 | 10,0% | 4 | 20,0% |
| Anemia | 1 | 5,0% | 2 | 10,0% |
| ALT/AST increase | 1 | 5,0% | 0 | 0,0% |
| Creatinine increase | 1 | 5,0% | 0 | 0,0% |
| Thrombosis | 1 | 5,0% | 1 | 5,0% |
| Intestinal obstruction | 0 | 0,0% | 1 | 5,0% |
| Skin rash G1 | 0 | 0,0% | 2 | 10,0% |
| Itching G1 | 0 | 0,0% | 2 | 10,0% |

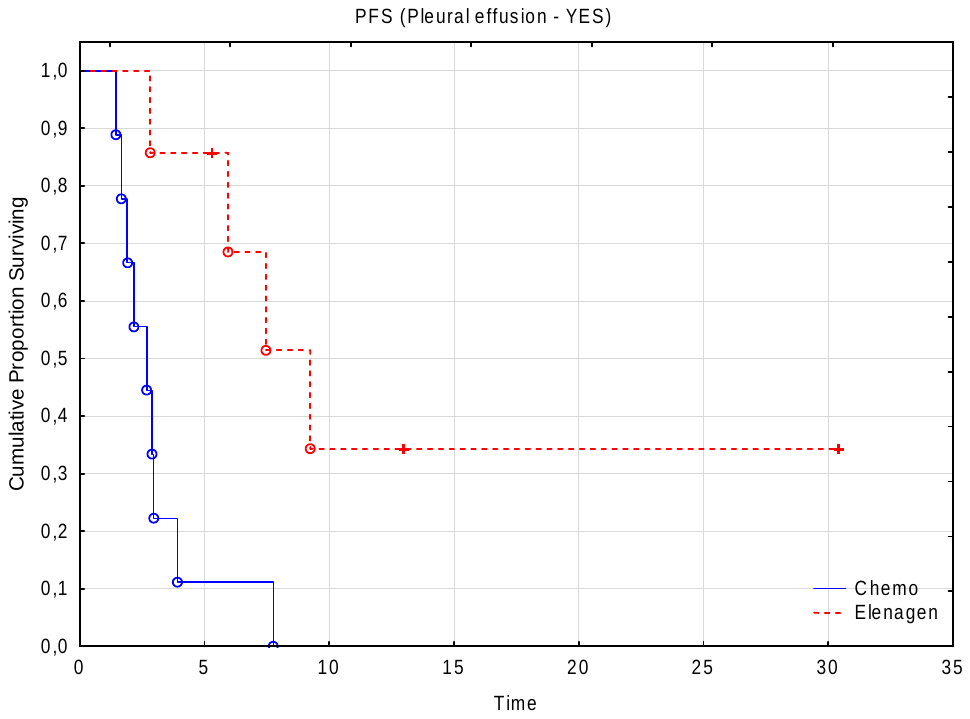

Median PFS of patients with peritoneal effusion in the groups.

| Percentiles | Chemo | ELENAGEN |
| --- | --- | --- |
| 25'th percentile (lower quartile) | 1,7 mo | 4,8 mo |
| 50'th percentile (median) | 2,4 mo | 7,6 mo |
| 75'th percentile (upper quartile) | 2,9 mo |  |
| P (log-Rank)=0.008 | | |

Suppl Fig. 1. Progression -free survival of patients treated with Chemo+ELENAGEN, or Chemo only in subgroups of patients with peritoneal effusion.
